# Does food-related cultural capital contribute to diet inequality in rural Australia? A pilot cross-sectional study

**DOI:** 10.1101/2020.10.22.20218040

**Authors:** Xiaozhou Zhang, Claudia Slimings

**Affiliations:** ANU Medical School, Australian National University, Australia; Rural Clinical School, ANU Medical School, Australian National University, Australia

**Keywords:** rural, diet, nutrition, cultural capital, social determinants, inequalities

## Abstract

**Objective:** Regional Australians have a poorer quality of diet compared to people living in metropolitan areas. Food-related cultural capital is one potential mechanism linking social determinants and nutrition. Relationships between food-related cultural capital and diet were investigated as an explanation for nutritional inequalities in regional New South Wales (NSW).

**Design:** A cross-sectional survey of rural NSW adults was conducted from 12^th^ October 2019 to 31^st^ March 2020 focusing on diet, socio-economic factors and cultural capital. Three dietary outcomes were assessed: fruit consumption, vegetable consumption, and a healthy diet score. Food-related cultural capital was analysed as objectivised and total incorporated cultural capital separately. The survey was distributed online with social media promotion.

**Setting:** Regional south-eastern and western NSW

**Participants:** 448 adults (median age 57 years) of whom 93% were female.

**Results:** In unadjusted analysis, both total incorporated and objectivised cultural capital had strong correlations with all three diet outcomes, with low cultural capital associated with poorer nutrition. After adjusting for sociodemographic confounders, low total incorporated cultural capital showed modest associations with low fruit consumption (OR = 1.69, 95%CI = 1.00 – 2.87), low vegetable consumption (OR = 2.94, 95%CI = 1.69 – 5.11) and low diet score (OR = 3.35, 95% CI = 1.59 – 4.71).

**Conclusions:** Food-related cultural capital, particularly incorporated cultural capital, was independently associated with healthy food consumption in regional NSW. This new finding presents potential avenues to improve the diet of rural Australians through diet-related education, promoting food participation and reducing barriers to healthy food access.

## Introduction

An unhealthy diet low fruit and vegetable consumption is a major preventable risk factor contributing to the burden of disease in Australia.^(1)^ In 2015, 7.3% of the total burden of disease was caused by low quality diet ^(2)^. In a 2017-18 report, less than 1 in 10 Australian adults met the daily recommended vegetable consumption.^(2)^ Poor diet can lead to health risk factors such as obesity, hypercholesterolaemia and hypertension, which can further contribute to the development of associated chronic conditions including type 2 diabetes and cardiovascular disease. These conditions are now the leading contributors to morbidity and mortality in Australia.^(3)^ Poor diet contributes greatly to rural health inequality. Rural Australians in general have poorer diet compared to their urban counterparts resulting in a higher prevalence of obesity and hence more significant burden from diet-related chronic diseases. ^(4–6)^ Diet-related risk factors are modifiable as dietary choices are influenced by underlying social, economic and environmental factors. A socioeconomic gradient is a major contributor to diet inequality in developed countries, which is only partially explained by the unequal distribution of social and economic resources.^(7)^ With this in mind, more recent studies have linked cultural resources to diet and health inequalities, and have identified cultural capital, constructed by culture-based activities, knowledge and perceptions, as a good predictor for diet quality.^(7–13)^ For example, a Dutch study reported that low cultural capital was strongly correlated with poor diet outcomes^(7)^ while a study in Norway reported cultural capital as a good predictor for healthy eating patterns amongst adolescents.^(8)^

Culture is a resource acquired through social learning that is critical to people’s behaviour, norms, values and knowledge.^(14)^ Culture has great impact on the type and amount of food that is considered adequate to a group of people, thus influencing their overall diet.^(7)^ Cultural capital, which enables social mobility of an individual in a society, was described by the French sociologist Pierre Bourdieu to have three states: incorporated cultural capital (e.g. values, skills, cultural participation), objectivised cultural capital (e.g. books, tools) and institutionalised cultural capital (e.g. educational degrees, professional titles).^(15)^ Bourdieu’s cultural capital theory has been further transformed into food-related cultural capital where the incorporated cultural capital includes food participation, food-related skills and nutrition knowledge, the objectivised cultural capital includes cooking equipment possession and institutionalised cultural capital includes education level of the family and individual.(^7, 9^) Amongst these, incorporated cultural capital has the largest potential to be the mediator between socioeconomic gradient and healthy dietary intake. Since the identification of food-related cultural capital, very few studies have been conducted to explore the impact of this factor on diet and nutrition. To the best of our knowledge, the relationship between food-related cultural capital and diet in regional Australia has not been studied.

The aim of this study is to examine associations between food-related cultural capital, specifically, incorporated cultural capital and objectivised cultural capital, and diet in adult residents of regional south-eastern and western areas of New South Wales (NSW), Australia.

## Methods

### Participants

An online survey was conducted targeting adults who live in the south-eastern and western geographic area of NSW, Australia, covered by three primary healthcare networks (PHN): South Eastern NSW PHN, Murrumbidgee PHN and Western NSW PHN. Volunteer participants were recruited through a Facebook promotion program that ran from 12^th^ October 2019 to 31^st^ March 2020. The survey questionnaire was developed using Qualtrics (© 2019 Qualtrics LLC) and data was exported from Qualtrics to SPSS (version 24) for analysis. This project was approved by the ANU Ethics Committee (Protocol 2019/394). This study is reported according to the Strengthening the Reporting of Observational Studies in Epidemiology (STROBE) statement.^(16)^

### Data collection

The questionnaire used previously validated questions from existing surveys. The main source of questions was a large study conducted in the Netherlands examining the correlation between dietary outcome and food-related cultural capital ^(7)^, which was translated to Australian setting based on National Health and Medical Research Council (NHMRC) Australian Dietary Guidelines.^(17)^ Questions on eating were adapted from the diet survey published by University of Newcastle.^(18)^ The full list of questions used in the survey are presented in Supplementary Table 1. The validity of the questions was further checked through consultation with academic colleagues with experience in rural health research.

**Table 1.** Data characteristics and associations between demographic, socioeconomic factors and food-related cultural capital.

|  | Total, n (%)<sup>†</sup> | Objectivised cultural capital, n (%)<sup>‡</sup> |  |  |  | Total incorporated cultural capital, n (%)<sup>§</sup> |  |  |  |
| --- | --- | --- | --- | --- | --- | --- | --- | --- | --- |
|  |  | Low | Medium | High | p-value | Low | Medium | High | p-value |
| **Gender** |  |  |  |  |  |  |  |  |  |
| Female | 416 (92.9) | 43 (11.0) | 163 (41.6) | 186 (47.4) | 0.50 | 142 (40.0) | 59 (16.6) | 154 (43.3) | 0.87 |
| Male | 28 (6.3) | 4 (16.0) | 10 (40.0) | 11 (44.0) |  | 12 (48.0) | 4 (16.0) | 9 (36.0) |  |
| Other | 3 (0.7) | 1 (33.3) | 0 (0) | 2 (66.7) |  | 1 (33.3) | 1 (33.3) | 1 (33.3) |  |
| Missing | 1 (0.2) | 0 | 1 (0.6) | 0 |  | 0 | 1 (1.5) | 0 |  |
| **Age** |  |  |  |  |  |  |  |  |  |
| 18-35 | 32 (7.1) | 11 (36.7) | 16 (53.3) | 3 (10.0) | < 0.01 | 18 (64.3) | 3 (10.7) | 7 (25.0) | <0.01 |
| 36-55 | 168 (37.5) | 14 (9.0) | 72 (46.2) | 70 (44.9) |  | 73 (49.7) | 21 (14.3) | 53 (36.1) |  |
| 56-69 | 189 (42.2) | 19 (10.6) | 69 (38.3) | 92 (51.1) |  | 51 (31.3) | 30 (18.4) | 82 (50.3) |  |
| >= 70 | 55 (12.3) | 4 (7.8) | 16 (31.4) | 31 (60.8) |  | 11 (25.6) | 11 (25.6) | 21 (48.8) |  |
| Missing | 4 (0.9) | 0 | 0 | 3 (1.5) |  | 0 | 0 | 1 (0.6) |  |
| **Remoteness** |  |  |  |  |  |  |  |  |  |
| Inner regional | 208 (46.1) | 21 (10.9) | 76 (39.4) | 96 (49.7) | 0.06 | 63 (36.2) | 31 (17.8) | 81 (46.0) | 0.55 |
| Outer regional or remote | 240 (53.6)) | 27 (11.9) | 96 (42.5) | 103 (45.6) |  | 92 (44.0) | 34 (16.3) | 83 (39.7) |  |
| Missing | 0 (0) | 0 | 0 | 0 |  | 0 | 0 | 0 |  |
| **Area-level quartile of SEP\*** |  |  |  |  |  |  |  |  |  |
| 1 (most disadvantaged) | 183 (40.8) | 26 (15.1) | 73 (42.4) | 73 (42.4) | 0.30 | 70 (44.0) | 30 (18.9) | 59 (37.1) | 0.10 |
| 2 | 52 (11.6) | 5 (9.8) | 19 (37.3) | 27 (52.9) |  | 20 (41.7) | 9 (18.8) | 19 (39.6) |  |
| 3 | 108 (24.1) | 9 (9.2) | 39 (39.8) | 50 (51.0) |  | 40 (44.4) | 14 (15.6) | 36 (40.0) |  |
| 4 (least disadvantaged) | 103 (23.0) | 6 (6.1) | 43 (43.9) | 49 (50.0) |  | 25 (29.4) | 11 (12.9) | 49 (57.6) |  |
| Missing | 2 (0.4) | 1 (2.1) | 0 | 0 |  | 0 | 1 (1.5) | 1 (0.6) |  |
| **Employment status** |  |  |  |  |  |  |  |  |  |
| Employed | 265 (59.2) | 31 (12.4) | 111 (44.2) | 109 (43.4) | <0.01 | 100 (42.7) | 35 (15.0) | 99 (42.3) | 0.33 |
| Retired | 113 (25.2) | 5 (4.7) | 5 (33.0) | 66 (62.3) |  | 30 (33.0) | 17 (18.7) | 44 (48.4) |  |
| Other inactive | 70 (15.6) | 12 (18.8) | 28 (43.8) | 24 (37.5) |  | 25 (42.4) | 13 (22.0) | 21 (35.6) |  |
| Missing | 0 (0) | 0 | 0 | 0 |  | 0 | 0 | 0 |  |
| **Education level** |  |  |  |  | 0.47 |  |  |  | <0.01 |
| Postgraduate degree | 150 (33.5) | 14 (9.7) | 58 (40.3) | 72 (50.0) |  | 42 (33.6) | 22 (17.6) | 61 (48.8) |  |
| Bachelor degree | 151 (33.7) | 23 (16.0) | 57 (39.6) | 64 (44.4) |  | 46 (33.6) | 17 (12.4) | 74 (54.0) |  |
| Finished high school or equivalent | 130 (29.0) | 10 (8.5) | 51 (43.6) | 56 (47.9) |  | 61 (57.5) | 20 (18.9) | 25 (23.6) |  |
| Did not finish high school | 16 (3.6) | 1 (6.7) | 8 (53.3) | 6 (40.0) |  | 6 (40.0) | 6 (40.0) | 3 (20.0) |  |
| Missing | 1 (0.2) | 0 | 0 | 1 (0.5) |  | 0 | 0 | 1 (0.6) |  |
| **Annual household income** |  |  |  |  | 0.48 |  |  |  | 0.63 |
| Less than \$20,000 | 43 (9.6) | 2 (5.1) | 17 (43.6) | 20 (51.3) | | 16 (45.7) | 8 (22.9) | 11 (31.4) | |
| \$20,001 - \$40,000 | 66 (14.7) | 10 (16.1) | 23 (37.1) | 29 (46.8) | | 19 (34.5) | 11 (20.0) | 25 (45.5) | |
| \$40,001 - \$60,000 | 82 (18.3) | 12 (15.0) | 29 (36.3) | 39 (48.8) | | 29 (39.2) | 10 (13.5) | 35 (47.3) | |
| \$60,001 - \$80,000 | 36 (8.0) | 3 (8.3) | 17 (47.2) | 16 (44.4) | | 15 (50.0) | 2 (6.7) | 12 (43.3) | |
| \$80,001 - \$100,000 | 59 (13.2) | 6 (10.3) | 28 (48.3) | 24 (41.4) | | 25 (46.3) | 9 (16.7) | 20 (37.0) | |
| \$100,001 - \$150,000 | 87 (19.4) | 7 (8.2) | 42 (49.4) | 36 (42.4) | | 30 (37.5) | 11 (13.8) | 39 (48.8) | |
| More than \$150,000 | 54 (12.1) | 5 (9.4) | 17 (32.1) | 31 (58.5) | | 17 (35.4) | 11 (22.9) | 20 (41.7) | |
| Missing | 21 (4.7) | 3 (6.1) | 2 (1.1) | 4 (2.0) |  | 4 (2.6) | 3 (4.6) | 2 (1.2) |  |
| **Barrier to access of healthy food** |  |  |  |  | 0.87 |  |  |  | 0.01 |
| Never/rarely | 333 (74.3) | 39 (11.9) | 139 (42.5) | 149 (45.6) |  | 114 (36.8) | 54 (17.4) | 142 (45.8) |  |
| Sometimes/often/always | 61 (13.6) | 7 (11.9) | 23 (39.0) | 29 (49.2) |  | 32 (58.2) | 6 (10.9) | 17 (30.9) |  |
| Missing | 54 (12.1) | 2 (4.2) | 12 (6.9) | 21 (10.6) |  | 9 (5.8) | 5 (7.7) | 5 (3.0) |  |
| **Vegetarian** |  |  |  |  | 0.58 |  |  |  | 0.36 |
| Yes | 36 (8.0) | 5 (13.9) | 12 (33.3) | 19 (52.8) |  | 10 (32.3) | 4 (12.9) | 17 (54.8) |  |
| No | 399 (89.1) | 43 (11.2) | 162 (42.1) | 180 (46.8) |  | 145 (41.1) | 61 (17.3) | 137 (41.6) |  |
| Missing | 13 (2.9) | 0 | 0 | 0 |  | 0 | 0 | 0 |  |
| **Frequency of eating out or eating take-away** |  |  |  |  |  |  |  |  |  |
| Twice or more weekly | 83 (18.5) | 20 (24.7) | 34 (42.0) | 27 (33.3) | <0.01 | 27 (37.0) | 15 (20.5) | 31 (42.5) | 0.62 |
| Once or less weekly | 352 (78.6) | 28 (8.2) | 140 (41.2) | 172 (50.6) |  | 128 (41.2) | 50 (16.1) | 133 (42.8) |  |
| Missing | 13 (2.9) | 0 | 0 | 0 |  | 0 | 0 | 0 |  |

\*SEP, socioeconomic position
<sup>†</sup>Percentage of the total of 448 participants. All other represent are the percentage in each category of predictor variable.
<sup>\*</sup>Total number of participants with data on objectivised cultural capital was 421.
<sup>§</sup>Total number of participants with data on incorporated cultural capital was 384.

#### Dietary outcome measures

Participants were asked four questions related to fruit, vegetable, protein, and bread consumption. Three dietary outcome variables were analysed: daily fruit consumption, weekly vegetable consumption, and a healthy diet score derived from responses to all four questions. In consideration of the difficulty for survey respondents to calculate dietary intake in cups, fruit consumption was recorded as pieces per day (2 or more, 1 or less) and vegetable consumption was recorded as times per week (1 or less, 2 – 3, 4 – 5, and 6 or more) and analysed as binary variables to approximate whether the recommended dietary guidelines for fruit and vegetable consumption were met according to the Australian Dietary Guidelines.^(17)^ The survey equivalent for meeting the guidelines for fruit consumption was defined as 2 or more pieces per day and for vegetable consumption was defined as 6 or more times per week.

The healthy diet score was derived from the sum of the scores in fruit consumption, vegetable consumption, protein consumption (how many times per week meat, fish, seafood, nuts, beans, eggs or tofu are consumed with a meal) and usual bread consumption (none; brown/multigrain/wholemeal; white; other). For each of these variables, a score of 1 was given to the unhealthiest option while the highest score was given to the healthiest option. The possible minimum diet score was 4 and the maximum was 14. The diet score was then converted to a binary variable (low diet score = 5 – 11, high diet score = 12 – 14) based on the median value of the score.

#### Food-related cultural capital indicators

The derivation of these indicators followed the same methodology used in the original Netherlands study.^(7)^ Objectivised food-related cultural capital was measured by possession of specific cooking objects, namely a stove, cookbook(s), set of knives, kitchen scale, and juicer. A sum score for total possessions owned (range 0-5) was created, which was divided into tertiles representing low (<4), medium (4) and high (5) objectivised capital.

Total incorporated cultural capital score was derived from participation in cultural activities, skills, and knowledge. Participation in activities was measured with two open-ended items, where participants indicated a frequency, namely: “In the last month, how many times have you met with people in a public place to have some food?” and “In the last month, how many times have people visited you in your home to have dinner, or have you visited people for dinner in their home?”. Responses to both questions were summed in one variable and divided into quintiles, with 1= low and 5= high food-related participation. Food choice-related skills were represented by cooking skills, grocery shopping skills, and skills to find and process information about nutrients and food preparation. Again, responses were summed and divided into quintiles, with 1= low skill level and 5= high skill level. Nutrition knowledge was assessed using four separate questions (Supplementary Table 1, Q18-22) that were each asked with regard to four different food products. The scores of all correct answers were summed (ranging from 0-16) and divided into quintiles, with 1=low and 5=high nutrition knowledge. Total incorporated cultural capital was calculated from the mean score of the participation, skills and knowledge variables and categorised into tertiles representing low (<7), medium (8-9) and high levels (>10).

### Covariates

#### Demographic variables

Survey participants reported their gender (male, female, other, prefer not to say), age (in years), and their full address of residence. Gender was analysed using a binary variable of female and non-female, due to the small number of male responders and participants with chose ‘other’ or ‘preferred not to say’ options. Residential addresses were geocoded to statistical area level 1 (SA1) and classified according to the Australian Statistical Geography Standard (ASGS) 2011 remoteness structure as Major Cities, Inner Regional, Outer Regional, Remote and Very Remote.^(19)^ Those classified as residents in major cities were excluded from analysis (n = 8). Due to few (n = 7) participants living in remote areas, data from remote and outer regional) areas were combined into one category, Outer Regional/Remote. This new binary variable of Inner Regional and Outer Regional/Remote was used for all subsequent analyses.

#### Social and economic indicators

Information on annual household income was recorded in the survey as seven ordered categories, and analysed as a continuous variable in subsequent statistical analyses where a linear relationship with diet outcomes was evident. Neighbourhood socioeconomic position (SEP) was assigned using the Index of Relative Socioeconomic Disadvantage, one of the Socio-economic Indexes for Areas (SEIFA), obtained from self-reported residential suburb of participants.^(20)^ The SEIFA indicates the socioeconomic characteristics of people living in an area, and provides a score of 1 – 10 for each area, with 1 being the lowest 10% of socio-economic index in the whole Australia and 10 being the top 10%. SEP was analysed as quintiles (i.e. the 1^st^ quintile represents individuals living in areas in the bottom 20% of the SEIFA); the top two quintiles were combined for analysis due to small group sizes. Self-reported employment status was recorded as full-time employee, part-time employee, self-employed, student, home duties, volunteer work only, unable to work, unemployed, and retired. Employment was recoded into three categories, namely employed (full time employee, part time employee, or self-employed), retired, and economically inactive (student, home duties, volunteer work only, unable to work, or unemployed) due to the small number of participants that fell into each specific category. The highest level of education obtained was collected and analysed with four categories: postgraduate degree, bachelor’s degree, finished high school or equivalent, and did not finish high school.

#### Eating habits and barrier to access of healthy food

Self-reported eating habits including vegetarianism (excluding pescatarian) and the frequency of eating out or eating take-away per week (twice or more or once or less) were also considered as potential confounding variables with strong association with diet choices. Barrier to access of healthy food was derived from the sum of scores of eight statements (Supplementary Table 1, Q25) using methodology reported previously.^(21)^

### Statistical analysis

For descriptive purposes, the mean, median, standard deviation (SD) and range were calculated for numerical variables; counts and percentages for categorical variables. To test whether the demographic, social and economic factors were associated with the three cultural capital variables, the chi-squared test (Fishers Exact if expected count less than 5) was used. Logistic regression was used to investigate the bivariate associations between the exposure, covariates and the binary dietary outcome variables (fruit consumption, vegetable consumption, diet score) reported as odds ratios (OR) and 95% confidence intervals (95% CI). Multiple logistic regression was used to investigate whether there was an independent association between food-related cultural capital and each dietary outcome. Specifically, covariate variables that were associated with the dependent variables and each of the cultural capital variables at p < 0.2 in bivariate analyses were considered confounders and selected for inclusion in multiple logistic regression models. In addition, employment and education levels were included regardless of bivariate relationships as they were considered important confounders based on existing literature.

## Results

### Demographic and socioeconomic characteristics

A total of 456 people attempted the online questionnaire, with 412 respondents (91%) completing all questions and 39 partial completion; 8 non-rural respondents were excluded leaving 448 participants. Of these, 93% were females, and the median age was 57 years (range 18 – 90 years). Forty-six percent of respondents resided in Inner Regional areas of NSW and only 7 respondents resided in Remote areas (Table 1). In addition, 41% of all respondents lived in areas with a neighborhood IRDS that was in the bottom 20^th^ centile Australia-wide. A large proportion of respondents were economically inactive (31%), and 62% of these were retired. A significant portion (34%) of respondents were highly qualified with a postgraduate degree. A higher number of respondents had self-reported annual household income over $150,000 (12%) compared to income less than $20,000 (10%). Most respondents (74%) had never or rarely experienced a barrier to access of healthy food. A small percentage (8%) of respondents were vegetarians and 18.5% of all respondents ate out or take-aways at least twice weekly.

### Relationships between demographic and socioeconomic characteristics and cultural capital predictors

Objectivised cultural capital score was computed for 421 respondents (94%). Overall, 44.4% of respondents, had high objectivized cultural capital scores, 38.8% had medium scores, 10.7% had low scores. The level of objectivised cultural capital was highest in older and retired individuals, and people who ate out or had take-aways less often (Table 1). There was a small difference for level of remoteness, with 49.7% in inner regional having high levels compared to 45.6% in outer regional/remote areas.

Out of 448 respondents, 384 total incorporated cultural capital scores were calculated (85.7%). Overall, 34.6% of respondents had low incorporated cultural capital scores compared to 14.5% and 36.6% of individuals who had medium and high scores respectively. Increased age, and higher education level, and never or rarely experiencing barriers to access to healthy food were associated with higher total incorporated cultural capital (Table 1).

### Relationships between socioeconomic factors, cultural capital, dietary habits and dietary outcomes

In general, 51.3% of respondents failed to meet the recommended daily fruit consumption and 45.3% failed to meet the recommended vegetable consumption guidelines. A diet score was calculated for 433 respondents. The mean and median of diet score were 11.4 and 12.0, respectively (SD = 2.03, range = 5 - 14). This was then transformed into a binary variable containing 52.4% of respondents in the high diet score category (12 - 14) and 47.6% in the low score category (Supplementary Figure 1).

The associations for the three dietary outcomes are presented in Tables 2 to 4. In general, low food-related cultural capital was related to increased odds of having poorer diet measured by the three outcome variables. Total incorporated cultural capital was more consistently and strongly associated with the dietary outcomes compared to objectivized cultural capital.

**Table 2.**
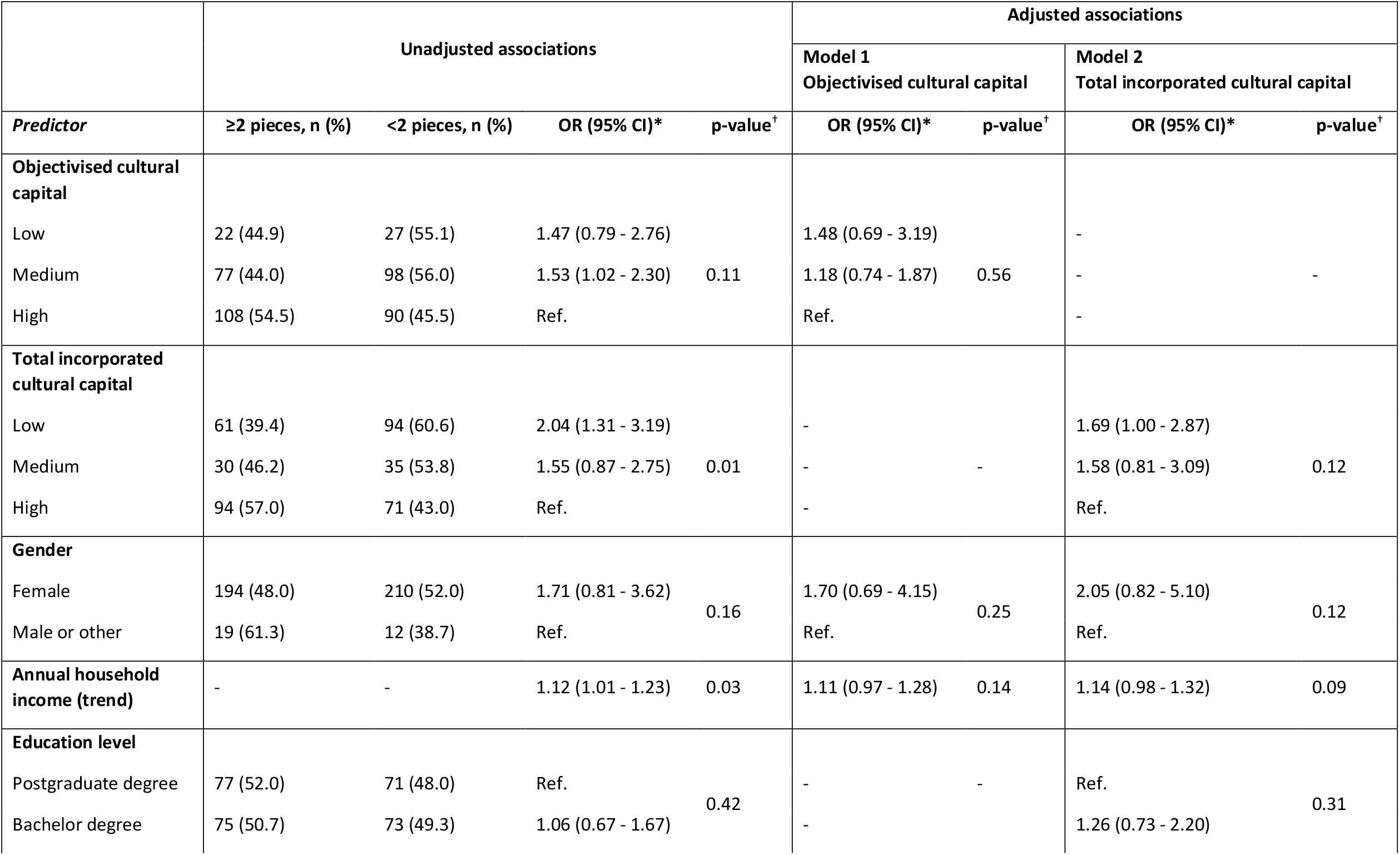
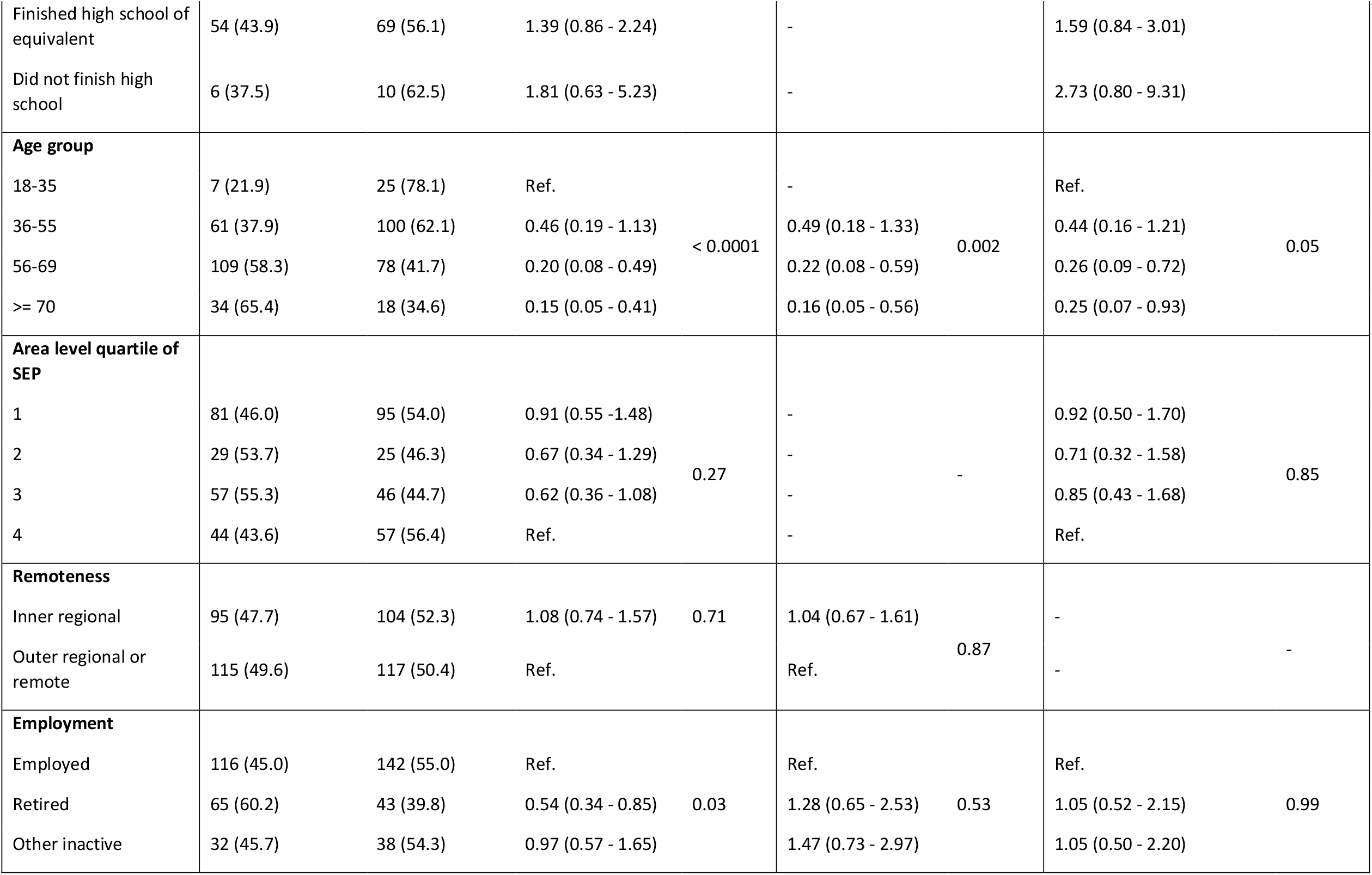
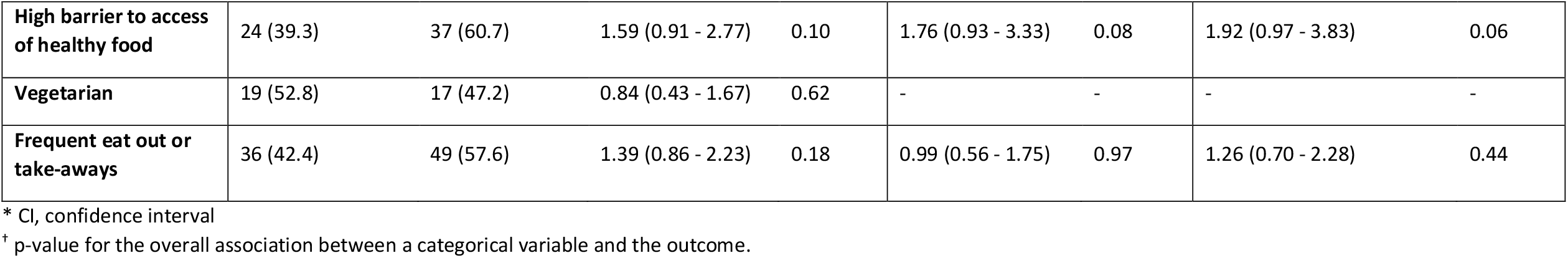
Associations between food-related cultural capital and low fruit consumption.

In bivariate analyses, low fruit consumption was associated with both low objectivised cultural capital (OR = 1.47, 95%CI = 0.79 – 2.76) and low incorporated cultural capital (OR = 2.04, 95%CI = 1.31 – 3.19) compared to those with a high score on each predictor, although the overall association between objectivised cultural capital was not statistically significant (Table 2). In multivariate analysis, associations for both cultural capital predictors were attenuated by a small amount. Specifically, the odds of having low fruit consumption was 1.69 times higher for the group with low total incorporated cultural capital compared to the group with high score (95%CI = 1.00 – 2.87), whereas the odds ratio for low objectivised cultural capital was 1.48 (95%CI = 0.69 – 3.19).

Low vegetable consumption had a stronger association with low compared to high total incorporated cultural capital (OR = 3.48, 95%CI = 2.19 – 5.51) than for low objectivised cultural capital (OR = 1.49, 95% CI = 0.79 – 2.78) (Table 3). After adjustment for covariates including income, education level, SEP, being a vegetarian and frequent eating out or eating take-aways, the association between low vegetable consumption and low total incorporated cultural capital remained significant (OR = 2.94, 95%CI = 1.69 – 5.11). On the other hand, the association for objectivised cultural capital was substantially attenuated.

**Table 3.** Associations between food-related cultural capital and low vegetable consumption.

|  | Unadjusted |  |  |  | Adjusted |  |  |  |
| --- | --- | --- | --- | --- | --- | --- | --- | --- |
|  |  |  |  |  | Model 1<br>Objectivised cultural capital |  | Model 2<br>Total incorporated cultural capital |  |
| *Predictor* | ≥6 times per week, n (%) | <6 times per week, n (%) | OR (95% CI)* | p-value<sup>†</sup> | OR (95% CI)* | p-value<sup>†</sup> | OR (95% CI)* | p-value<sup>†</sup> |
| **Objectivised cultural capital** |  |  |  |  |  |  |  |  |
| Low | 24 (49.0) | 25 (51.0) | 1.49 (0.79 - 2.78) | 0.13 | 1.18 (0.56 - 2.49) | 0.36 | - | - |
| Medium | 86 (48.9) | 90 (51.1) | 1.49 (0.99 - 2.25) |  | 1.39 (0.89 - 2.17) |  | - |  |
| High | 117 (58.8) | 82 (41.2) | Ref. |  | Ref. |  | - |  |
| **Total incorporated cultural capital** |  |  |  |  |  |  |  |  |
| Low | 60 (38.5) | 96 (61.5) | 3.48 (2.19 - 5.51) | < 0.01 | - | - | 2.94 (1.69 - 5.11) | <0.01 |
| Medium | 33 (50.8) | 32 (49.2) | 2.11 (1.17 - 3.79) |  | - | - | 1.56 (0.80 - 3.07) |  |
| High | 113 (68.5) | 52 (31.5) | Ref. |  | - | - | Ref. |  |
| **Gender** |  |  |  |  |  |  |  |  |
| Female | 216 (53.2) | 190 (46.8) | 0.94 (0.45 - 1.95) | 0.86 | - | - | - | - |
| Male or other | 15 (46.9) | 17 (53.1) | Ref. |  | - |  | - |  |
| **Annual household income (trend)** | - | - | 0.91 (0.82 - 1.00) | 0.05 | 0.90 (0.78 - 1.03) | 0.13 | 0.95 (0.82 - 1.09) | 0.44 |
| **Education level** |  |  |  |  |  |  |  |  |
| Postgraduate degree | 87 (58.8) | 61 (41.2) | Ref. |  | Ref. |  | Ref. |  |
| Bachelor degree | 88 (59.1) | 61 (40.9) | 0.99 (0.62 - 1.57) |  | 0.93 (0.56 - 1.55) |  | 0.98 (0.56 - 1.74) |  |
| Finished high school of equivalent | 51 (41.1) | 73 (58.9) | 2.04 (1.26 - 3.32) | 0.006 | 1.85 (1.06 - 3.23) | 0.055 | 1.27 (0.67 - 2.40) | 0.64 |
| Did not finish high school | 6 (37.5) | 10 (62.5) | 2.38 (0.82 - 6.89) |  | 1.86 (0.60 - 5.83) |  | 2.08 (0.60 - 7.18) |  |
| **Age group** |  |  |  |  |  |  |  |  |
| 18-35 | 15 (46.9) | 17 (53.1) | Ref. |  | Ref. |  | Ref. |  |
| 36-55 | 85 (52.5) | 77 (47.5) | 0.80 (0.37 - 1.71) |  | 1.36 (0.53 - 3.47) |  | 1.39 (0.56 - 3.46) |  |
| 56-69 | 106 (56.7) | 81 (43.3) | 0.67 (0.32 - 1.43) | 0.62 | 1.36 (0.53 - 3.50) | 0.60 | 1.00 (0.39 - 2.55) | 0.61 |
| >= 70 | 26 (49.1) | 27 (50.9) | 0.92 (0.38 - 2.21) |  | 2.10 (0.67 - 6.62) |  | 1.35 (0.42 - 4.29) |  |
| **Area level quartile of SEP** |  |  |  |  |  |  |  |  |
| 1 | 90 (50.6) | 88 (49.4) | 1.69 (1.03 - 2.79) |  | 1.23 (0.71 - 2.14) |  | 1.35 (0.71 - 2.54) |  |
| 2 | 25 (46.3) | 29 (53.7) | 2.01 (1.03 - 3.92) |  | 1.49 (0.72 - 3.09) |  | 1.68 (0.74 - 3.82) |  |
| 3 | 52 (50.5) | 51 (49.5) | 1.70 (0.97 - 2.97) | 0.11 | 1.64 (0.89 - 3.03) | 0.41 | 1.69 (0.83 - 3.42) | 0.46 |
| 4 | 64 (63.4) | 37 (36.6) | - |  | - |  | - |  |
| **Remoteness** |  |  |  |  |  |  |  |  |
| Inner regional | 111 (55.2) | 90 (44.8) | 0.87 (0.60 - 1.27) |  | 0.89 (0.58 - 1.37) |  | - |  |
| Outer regional or remote | 120 (51.7) | 112 (48.3) | Ref. | 0.47 | Ref. | 0.61 | - | - |
| **Employment** |  |  |  |  |  |  |  |  |
| Employed | 143 (55.2) | 116 (44.8) | - |  | - |  | - | - |
| Retired | 59 (54.1) | 50 (45.9) | 1.05 (0.67 - 1.64) | 0.26 | 0.73 (0.38 - 1.41) | 0.62 | - |  |
| Other inactive | 31 (44.3) | 39 (55.7) | 1.55 (0.91 - 2.64) |  | 0.96 (0.9 - 1.88) |  | - |  |
| **High barrier to access of healthy food** | 33 (54.1) | 28 (45.9) | 0.99 (0.57 - 1.71) | 0.97 | - | - | 0.54 (0.28 - 1.07) | 0.08 |
| **Vegetarian** | 27 (75.0) | 9 (25.0) | 0.35 (0.16 - 0.76) | 0.01 | 0.29 (0.12 - 0.70) | 0.01 | 0.16 (0.05 - 0.51) | <0.01 |
| **Frequent eat out or take-aways** | 30 (35.3) | 55 (64.7) | 2.48 (1.52 - 4.06) | <0.01 | 3.00 (1.71 - 5.26) | <0.01 | 3.47 (1.87 - 6.46) | < 0.01 |

\* CI, confidence interval
† p-value for the overall association between a categorical variable and the outcome.

A lower score in any of the two cultural capital variables was associated with an overall low diet score, particularly for low incorporated cultural capital (OR=3.35, 95% CI = 1.59 – 4.71) (Table 4). Female sex, income, education, age, SEP, barrier to access of healthy food, frequent eating out or eating take-aways were identified as confounders. In adjustment with confounders, both cultural capital predictors had slightly weakened associations with low diet score, however an independent association with low incorporated capital remained. The odds of having low diet score was 2.74 times for the group with low incorporated cultural capital compared to the group with high score (95%CI = 1.59 – 4.71).

**Table 4.**
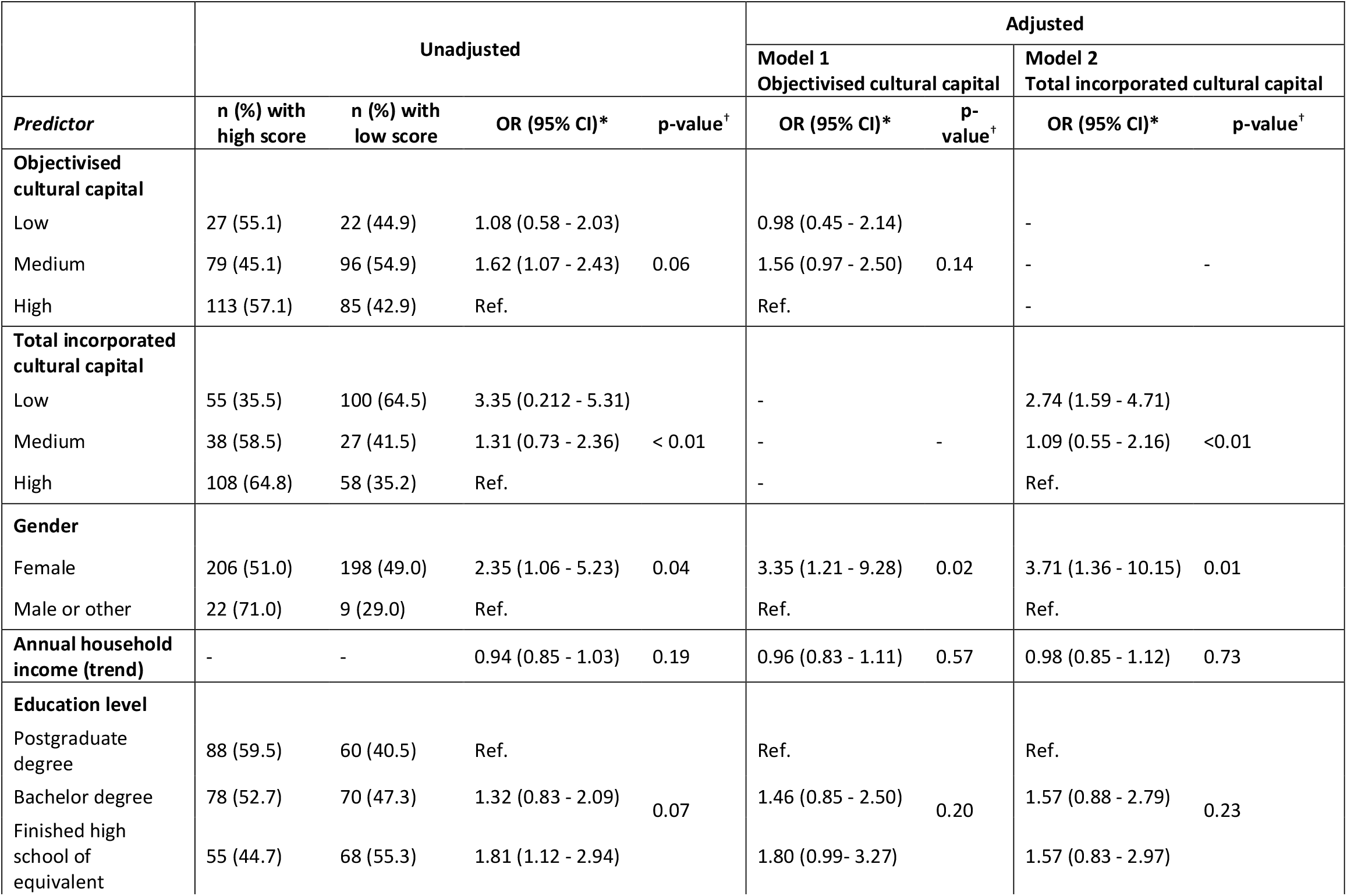
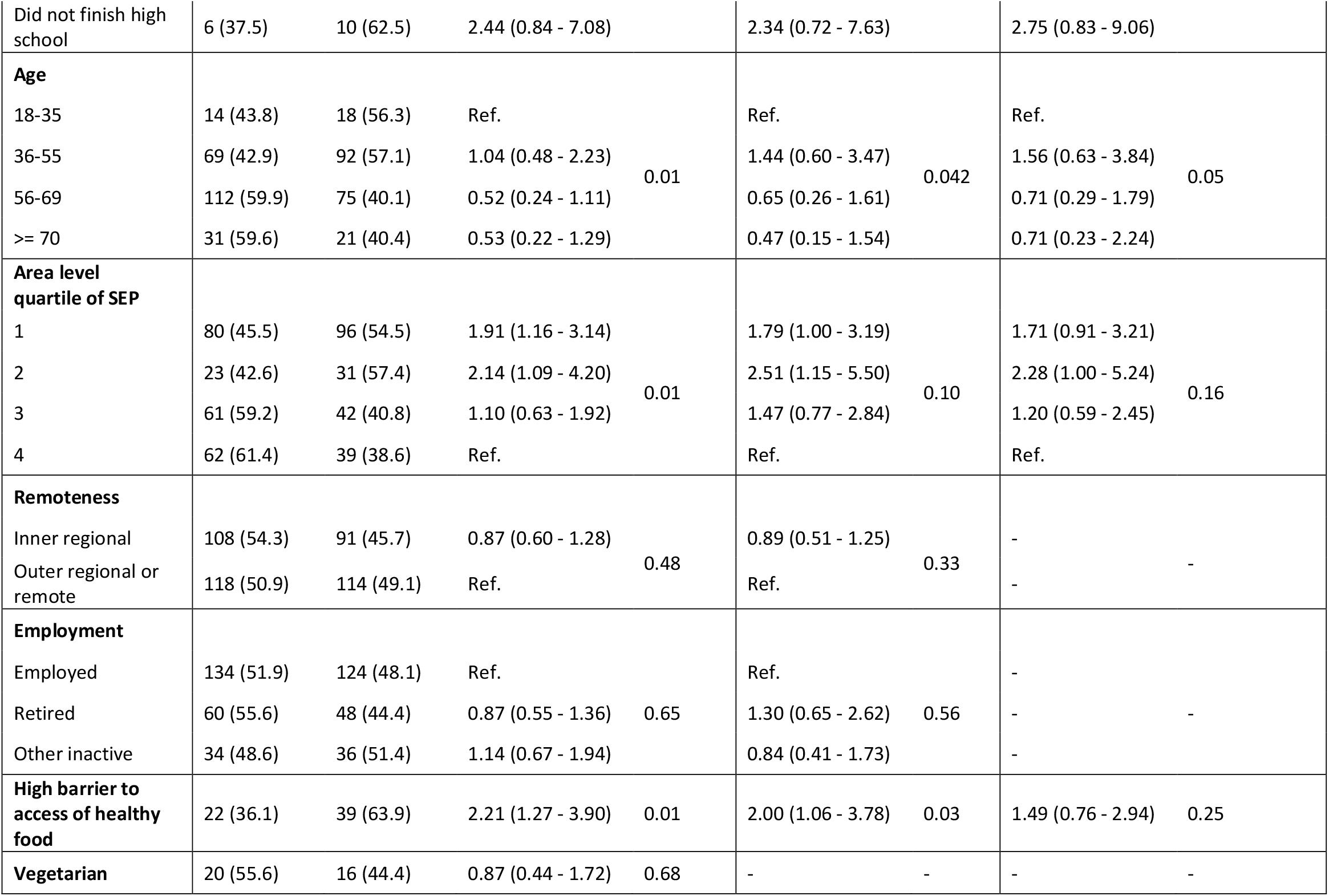
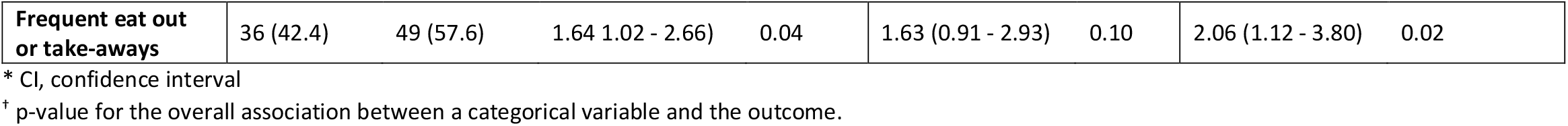
Associations between food-related cultural capital and low healthy diet score.

## Discussion

This is the first study to investigate the contributions of food-related cultural capital to diet quality, measured by fruit and vegetable consumption and a healthy diet score, among adults in rural Australia.

In this study of predominantly older, female residents of rural NSW, higher incorporated cultural capital measured by nutrition knowledge, food participation, and various food-related skills was found to be independently associated with a healthier diet after accounting for a range of sociodemographic confounders. Objectivised cultural capital measured by possession of cooking objects showed similar patterns of relationships with diet, however, the strength of the associations were weaker and firm conclusions could not be drawn.

Our findings that low food-related cultural capital was associated with low fruit and vegetable consumption and overall diet quality was consistent with previous literature.(^7, 10, 12^) As anticipated, total incorporated cultural capital is a stronger predictor for diet compared to objectivised cultural capital. This potentially relates to the method of calculation for these two variables where the total incorporated cultural capital included a much wider scope of factors compared to objectivised cultural capital. Cultural capital is related to a number of social determinants of health, namely income, socio-economic status, cultural practices, education, food access, and social interactions.^(15)^ Cultural capital affects the conscious choice of health promoting behaviour through a complex interplay with the operational skills, knowledge, health considerations and norms acquired through social interactions and education, thus influencing diet outcomes.^(14)^ For example, people who own more pieces of cooking equipment (high objectivised cultural capital) and possess wider diet-related knowledge (high incorporated cultural capital) are more likely to make a conscious choice to have a diversified and healthy diet. Thus, cultural capital links structural and behavioural determinants of health by explaining how people’s behavioural options and preferences are related to lifestyle.^(22)^ Assuming a causal relationship between cultural capital and diet quality, improving cultural capital in regional Australian population can minimise the gap in diet and nutrition between groups with different socio-economic status. This can be achieved, at a community level, by promoting healthy eating patterns, optimising access to food related and nutritional information, holding frequent cooking classes and encouraging food participation.

Other than cultural capital, having difficulty accessing healthy food and frequently eating out or eating take-aways also demonstrated strong associations with poor diet as well as lower cultural capital. This is in line with previous literature that access to healthy food as well as cooking/eating at home is important for diet quality.(^21, 23, 24^) Education was used by several previous studies as a surrogate for the institutionalised cultural capital and was found to strongly impact diet quality.(^7, 9, 25^) This was also observed in our study. Compared to people with postgraduate degree, the risk of having poor diet doubles in participants who didn’t finish high school. In this study, being female was associated with a poor quality diet, which contradicts previous literature.^(26)^ This conflict can potentially be explained by the small sample size of non-female participants (7%), so the data distribution was significantly affected by selection bias.

As this was a pilot cross-sectional survey, causal conclusions cannot be made. This study has specific limitations to be addressed. The main limitation is selection bias because a volunteer sampling method was used, and the sample is therefore not representative of all rural NSW residents. The survey was conducted solely online, which may have limited participation from residents who lacked adequate internet access. Specifically, the study included adults who were English speakers, and the sample was 93% of female compared to 51% for non-metropolitan NSW, with a median age of 57 years (versus 43 years), and 31% were economically inactive (versus 6.6%).^(27)^ The study also had an over-representation of residents with high education level (bachelor degree and above, 67%) compared to 15% of residents in all of regional NSW.^(27)^

There was some missing data for total incorporated cultural capital scores because a large number of questions were used to deduce this score and thus participants were more likely to omit one or more related questions resulting in missing data. Due to the relatively small sample size, the results are subject to imprecision, as evidenced by wide confidence intervals and lack of statistical significance for some associations.

Accurate measurement of nutrition in large surveys presents specific challenges. In this study, 49% and 55% of respondents met the fruit and vegetable guidelines, respectively, which is comparable to the national estimate of 51.3% for fruit but substantially greater than the national estimate of 7.5% for vegetables.^(28)^ This could be explained by healthy volunteer bias where the volunteers that participated in this survey potentially pay more attention about their diet than the general population. This may also be due to the method of recording of vegetable and fruit consumption in the survey, which did not record portion sizes measured in cups as used in the NHMRC guidelines ^(17)^. These consumptions were measured in frequency or pieces to avoid difficulty of the participants to report their diet in cups. The results serve as an estimate to the participant’s vegetable and fruit consumption and guideline compliance instead of a precise measurement. Nevertheless, self-reported information may still be subject to recall bias. Despite the limitations in obtaining an accurate classification of participants as to whether they met dietary targets or not, this study demonstrates associations between relative levels of nutrition and food-related cultural capital.

This pilot cross-section population-based study is the first in rural Australia to examine new indicators of food-related cultural capital and diet quality. Larger, more representative studies using more precise measurements of diet are needed to confirm or refute the findings presented in this study.

## Conclusion

This study identifies a modest link between cultural capital and healthy food consumption in rural southeastern and western regions of NSW, Australia, that adds important new insights to the social determinants of nutrition. Cultural capital, particularly the incorporated cultural capital, was identified to be an important predictor of diet quality in addition to other major socioeconomic factors such as education and income. This new finding presents potential opportunities to improve the diet of rural Australians through diet-related education, promoting food participation and reducing barriers to healthy food access. However, further studies are required to confirm or refute these findings.

## Data Availability

Data will be archived following publication of outputs through the ANU Data Repository. Access to data requires permission of the data custodian.

