## Supplementary information for "Does food-related cultural capital contribute to diet inequality in rural Australia? A pilot cross-sectional study"

**Supplementary Table 1. Survey questions.**

| | |
|---|---|
| **General demographics** | |
| 1 | What is your age in years? |
| 2 | What it the gender that you identify yourself with? |
| 3 | What is your full residential address? |
| 4 | What is your highest educational qualification? |
| 5 | What is your marital status? |
| 6 | What is your employment status? |
| 7 | What is your total annual household income before tax or anything else is taken out? |
| **Dietary outcomes** | |
| 8 | About how many pieces of fruit do you eat each day? |
| 9 | About how many times a week do you eat vegetables with your main meal? Exclude hot chips. |
| 10 | How many times a week do you eat meat, fish, seafood, nuts, beans, eggs or tofu with your meal? |
| 11 | What type of bread do you usually eat? |
| **Objectivised cultural capital** | |
| 12a | Could you please indicate whether you own the following cooking objects? – Oven |
| 12b | Could you please indicate whether you own the following cooking objects? – Cookery books |
| 12c | Could you please indicate whether you own the following cooking objects? – Set of knives |
| 12d | Could you please indicate whether you own the following cooking objects? – Kitchen scales |
| 12e | Could you please indicate whether you own the following cooking objects? – Fruit juicer |
| **Total incorporated cultural capital** | |
| 13 | In the last month, how many times have you met with people in a public place to have some food? |
| 14 | In the last month, how many times have people visited your home to have dinner, or have you visited people for dinner in their home? |
| 15a | Below you may find two statements about grocery shopping. Please indicate for both statements how often this applies to you. – Before I go shopping, I make a list of everything I need. |
| 15b | Below you may find two statements about grocery shopping. Please indicate for both statements how often this applies to you. – Usually I do not decide what to buy until I am in the shop. |
| 16a | Below you may find three statements about cooking. Please indicate for each of the following statements whether you agree or disagree. – I know several ways to prepare fish. |
| 16b | Below you may find three statements about cooking. Please indicate for each of the following statements whether you agree or disagree. – I can prepare a low of meals even without a recipe. |
| 16c | Below you may find three statements about cooking. Please indicate for each of the following statements whether you agree or disagree. – I know several ways to prepare vegetables. |

| | |
|---|---|
| 17a | Below are questions about food information. Please indicate for each question how often this applies to you. – Do you read the nutrition information and information about ingredients on food packages? |
| 17b | Below are questions about food information. Please indicate for each question how often this applies to you. – Do you use the information about nutritional value on food packages to decide what foods you buy? |
| 17c | Below are questions about food information. Please indicate for each question how often this applies to you. – Do you look up information about foodstuffs on the internet? |
| 17d | Below are questions about food information. Please indicate for each question how often this applies to you. – Do you use recipes from cookery books, from the internet or from magazine? |
| 18a | Please indicate for the following four food items whether they are high or low in added sugar: - Bananas |
| 18b | Please indicate for the following four food items whether they are high or low in added sugar: - Unflavoured yogurt |
| 18c | Please indicate for the following four food items whether they are high or low in added sugar: - Ice-cream |
| 18d | Please indicate for the following four food items whether they are high or low in added sugar: - Tomato ketchup |
| 19a | Please indicate for the following four food items whether they are high or low in protein: - Chicken |
| 19b | Please indicate for the following four food items whether they are high or low in protein: - Cheese |
| 19c | Please indicate for the following four food items whether they are high or low in protein: - Fruit |
| 19d | Please indicate for the following four food items whether they are high or low in protein: - Broccoli |
| 20a | Please indicate for the following four food items whether they are high or low in fibre: - Eggs |
| 20b | Please indicate for the following four food items whether they are high or low in fibre: - Nuts |
| 20c | Please indicate for the following four food items whether they are high or low in fibre: - Chicken |
| 20d | Please indicate for the following four food items whether they are high or low in fibre: - Broccoli |
| 21a | Please indicate for the following four food items whether they are high or low in saturated fat: - Olive oil |
| 21b | Please indicate for the following four food items whether they are high or low in saturated fat: - Nuts |
| 21c | Please indicate for the following four food items whether they are high or low in saturated fat: - Red meat |
| 21d | Please indicate for the following four food items whether they are high or low in saturated fat: - Chocolate |
| **Other confounders - Vegetarian** | |
| 22 | Are you vegetarian? Please tick ‘No’ if you are pescatarian (eat fish or shellfish) |
| **Other confounders – Barriers to access of healthy food** | |
| 23 | How much time does it usually take to get to the grocery store for the major food shopping? |
| 24 | During the past 30 days, how much money did you spend at supermarkets or grocery stores? Please answer in Australian dollars. |

| | |
|---|---|
| 25a | How often do the following situations make it difficult for you/your household to get healthy foods? (healthy food includes fruits and vegetables, whole grains, beans and legumes, low-fat dairy, and lean meats)? – Distance to your usual grocery store |
| 25b | How often do the following situations make it difficult for you/your household to get healthy foods? (healthy food includes fruits and vegetables, whole grains, beans and legumes, low-fat dairy, and lean meats)? – Lack of transportation |
| 25c | How often do the following situations make it difficult for you/your household to get healthy foods? (healthy food includes fruits and vegetables, whole grains, beans and legumes, low-fat dairy, and lean meats)? – Hours your usual grocery store is open |
| 25d | How often do the following situations make it difficult for you/your household to get healthy foods? (healthy food includes fruits and vegetables, whole grains, beans and legumes, low-fat dairy, and lean meats)? – Price |
| 25e | How often do the following situations make it difficult for you/your household to get healthy foods? (healthy food includes fruits and vegetables, whole grains, beans and legumes, low-fat dairy, and lean meats)? – Physical disabilities |
| 25f | How often do the following situations make it difficult for you/your household to get healthy foods? (healthy food includes fruits and vegetables, whole grains, beans and legumes, low-fat dairy, and lean meats)? – Time available to go shopping |
| 25g | How often do the following situations make it difficult for you/your household to get healthy foods? (healthy food includes fruits and vegetables, whole grains, beans and legumes, low-fat dairy, and lean meats)? – Selection of items available at your usual grocery store |
| 25h | How often do the following situations make it difficult for you/your household to get healthy foods? (healthy food includes fruits and vegetables, whole grains, beans and legumes, low-fat dairy, and lean meats)? – Quality of items available at your usual grocery store |
| **Other confounders –Frequency of eating out or eating take-aways** | |
| 26 | How many times do you eat out/eat take-away food each week? |

**Supplementary Figure 1. Distribution and cut-offs for diet score.**

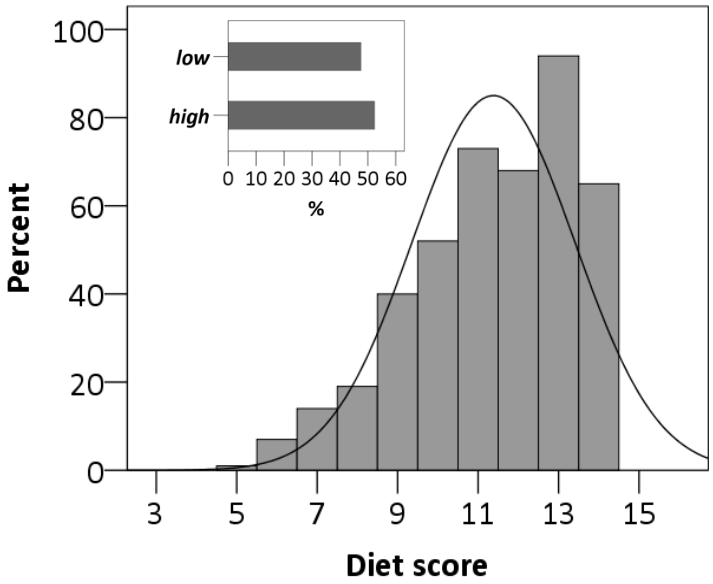
